# CHARACTERISTICS OF ACUTE PSYCHIATRIC DISORDERS IN PATIENTS WITH COVID-19 IN A THIRD-LEVEL HOSPITAL IN PERU

**DOI:** 10.1101/2023.02.16.23286046

**Authors:** Marcionila Estelita De La Cruz-Amador, Wilfor Aguirre-Quispe, Edwin Genaro Apaza-Aceituno, María Francesca Valdivia-Francia

## Abstract

**Objective:** To describe the sociodemographic and clinical characteristics of acute psychiatric disorders in COVID-19 patients in an emergency department at a national reference psychiatry and mental health hospital.

**Methods:** A descriptive observational study was performed. Data were collected from medical records of patients admitted by emergency according to the International Classification of Diseases (ICD-11). The group of patients with a first acute psychiatric episode vs. patients with more than one acute psychiatric episode were compared.

**Results:** 110 patients were included; 61.8% corresponded to the female sex and the mean age was 36 ± 12.3 years. 49.1% corresponded to schizophrenia, followed by acute polymorphic psychotic disorder (13.6%), bipolar disorder (10%), and depressive episodes (7.3%). Psychotic disorders and depressive episodes occurred in a higher percentage in the group with a first episode, 42.4% (p< 0.001), and 15.2% (p< 0.001), respectively. The episodes of schizophrenia were higher in the group of patients with previous episodes (63.6%).

**Conclusions:** A higher frequency of cases of acute psychotic disorder and depressive disorders was found as the first episode in patients with COVID-19 infection; however, within the group with previous episodes, greater predominance of patients with acute disorders due to schizophrenia was found.

## INTRODUCTION

More than 2 years have passed since the World Health Organization (WHO) declared coronavirus disease-19 (COVID-19) a pandemic on March 11, 2020, and despite time, we continue to learn about its effects^1^. Short-term consequences of COVID-19 have been described, but we are increasingly seeing more medium- and long-term effects, many still to be elucidated. Severe acute respiratory syndrome caused by Coronavirus 2 (SARS-CoV-2) is the etiological agent responsible for one of the most serious and far-reaching pandemics worldwide, with a multisystem involvement, from the respiratory system to various neuropsychiatric manifestations ^2,3^.

Due to its rapid transmission, on March 15, 2020, a state of national emergency was declared in Peru, and mandatory social isolation was established as a containment measure ^1,4^. The mandatory quarantine was proposed as a public health measure, with the intention of preventing the spread of COVID-19, however, it generated various stressors that negatively affected the mental health of the population, including: the prolonged duration of the quarantine, fear of infection, boredom and frustration, inadequate information, lack of supplies and the stigma towards people with COVID-19 infection ^5^. Consequently, the mental health problems that were most frequently observed in the population were anxiety, depression, insomnia, anger, fear, reaction to stress, and symptoms of posttraumatic stress disorder (PTSD) ^6^.

Over time, it has been possible to study the neuropsychiatric manifestations of COVID-19, but the true impact on the general population and vulnerable groups is still unknown, such as people with previous psychiatric disorders, who are at higher risk of contracting infections due to difficulty in understanding the magnitude of the problem, adhering to frequent hand washing and physical distancing ^2,7^. People with severe mental illnesses are more prone to the use of psychoactive substances and to present comorbidities such as obesity, diabetes mellitus, and cardiovascular diseases, worsening their prognosis once infected ^8–10^.

Therefore, people with a history of neuropsychiatric disorders are more vulnerable to severe illness, hospitalization, or death from COVID-19 ^8,9,11,12^. Specifically, people with schizophrenia, delusional disorders, and schizophreniform have higher mortality compared with those with mood disorders ^9,11,13^.

Psychiatric disorders can be a risk factor and a consequence of COVID-19 ^3^. Neuropsychiatric sequelae have been reported after SARS-CoV-2 infection, establishing an association between severe illness from COVID-19 and the manifestation of psychiatric symptoms ^14,15^. However, in post-COVID-19 follow-up studies, it has also been observed that people with mild or asymptomatic disease may present delirium, cognitive impairment, fatigue, and mood disturbances ^3,11,16^. The incidence of various disorders, such as insomnia, anxiety, suicidal ideation, confusion, and altered consciousness has also been reported ^2,7,13,17^.

This study describes the sociodemographic and clinical characteristics of acute psychiatric disorders in patients with COVID-19 in a hospital specializing in psychiatry and mental health, as well as to determine the most frequent mental disorders in this group of patients.

## METHODS

A retrospective observational study was conducted at the Hermilio Valdizan Psychiatric Hospital (HPHV), Lima, Peru. Data from medical and emergency records were collected during the period from May to December 2020.

The inclusion criteria considered patients admitted by the emergency service who were registered according to the main diagnosis code F00-F69 of the International Statistical Classification. of Diseases and Related Health Problems (ICD-11). These ICD codes include organic mental disorders, including symptomatic (F00-F09), and mental and behavioral disorders due to the use of psychoactive substances (F10-F19), schizophrenia, schizotypal and delusional disorders (F20-F29), mood [affective] (F30-F39), neurotic, stress-related, and somatoform disorders (F40-F48), behavioral syndromes associated with physiological disturbances and physical factors (F50-F59), and disorders of personality and behavior of the adult (F60-F69) and with COVID-19 infection confirmed by antigen test and/or PCR.

All included patients were classified according to the acute episode for which they went to the emergency: patients with a first episode vs. patients with ≥2 episodes. All the patients were evaluated by psychiatrists upon admission. This study was approved by The Hermilio Valdizan Hospital Research Ethics Committee. Because of the retrospective nature of the study, informed consent was not required to obtain the information since it was collected from the clinical histories and medical records. The data were stored in an anonymized database.

The sample was collected nonprobabilistically, and all patients who met the criteria within the specified period were included. The frequency values for each subgroup of psychiatric pathologies were calculated based on the total admissions.

The differences between groups, patients with a first episode vs patients with ≥2 episodes, were analyzed depending on the type of variable, using the chi2 test or Fisher’s exact test for the comparison of proportions and student t-test for comparison of means. Descriptive analysis is reported using means and standard deviation (SD) for quantitative variables, and proportions for qualitative variables.

All statistical tests were evaluated with a significance of 0.05 bilateral. The analyzes were performed with the STATA v.17.0 program (Texas, USA).

## RESULTS

A total of 110 emergency care records for the psychiatric diagnoses studied were included in the analysis. 61.8% corresponded to the female sex and the average age was 32 ± 12.3 years, the average BMI was 27.7 ± 5.5, 75% of the patients reported not having any type of occupation at the time of the evaluation, 74.5% reported being single, the predominant level of education was complete high school (65.5%). Regarding the history of previous hospitalizations, 44.5% reported having been previously hospitalized for a psychiatric diagnosis, all of them corresponding to the group with previous acute psychiatric episodes. The type of family was mainly nuclear and disintegrated, 33.6% in both cases. An unstable family dynamic was registered in 76.4%. Regarding the history, 95% reported not having presented a personal psychiatric history, while 36.4% presented a family psychiatric history. The use of psychoactive substances was registered in 23.6%, being mainly the consumption of alcohol, cocaine, marijuana, and tobacco. The general characteristics and by groups are shown in Table 1.

**Table 1:**
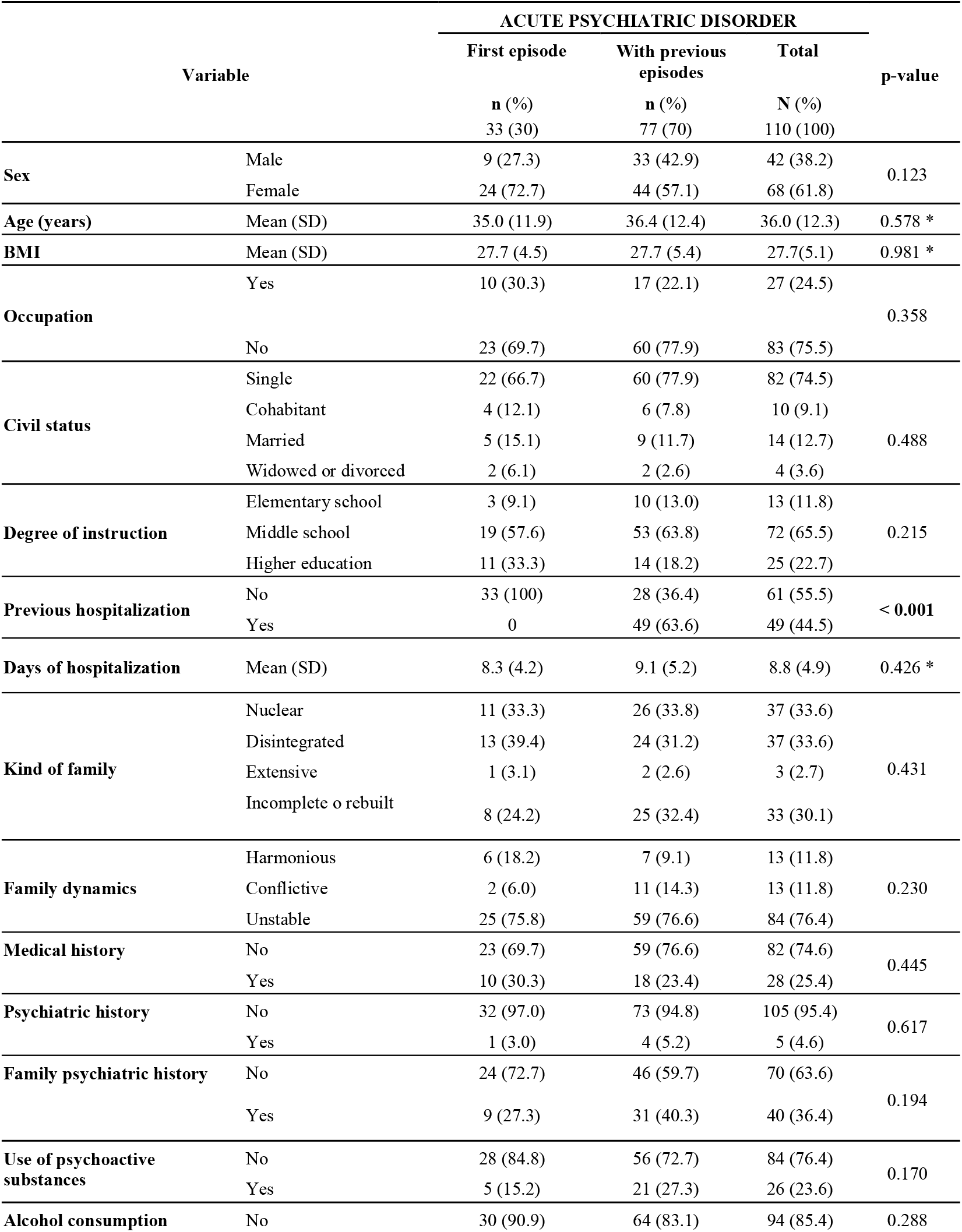
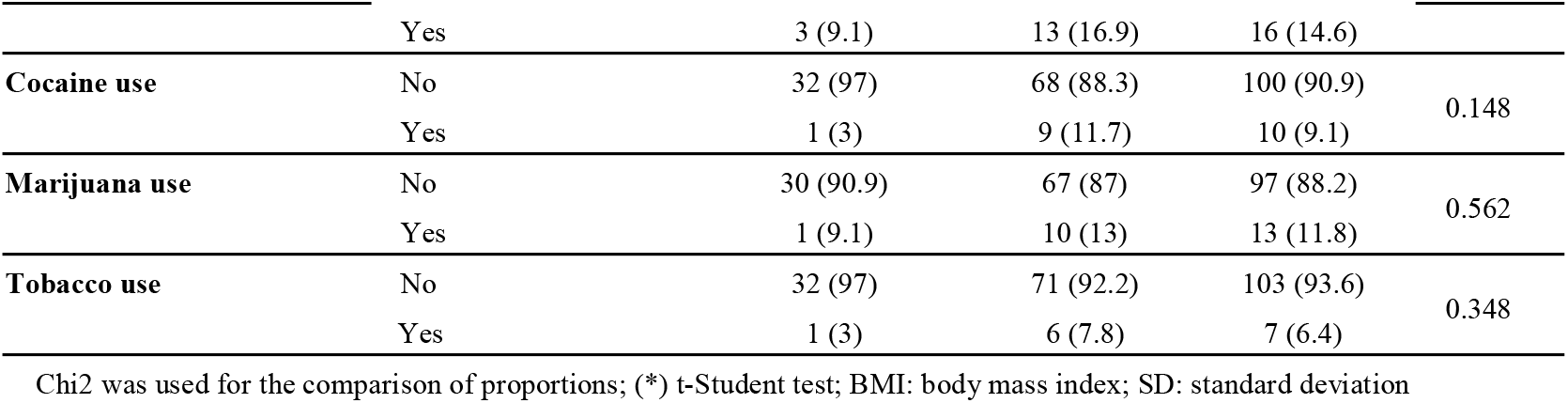
Sociodemographic and clinical characteristics according to the type of presentation of the acute psychiatric disorder.

In relation to psychiatric diagnoses, 49.1% corresponded to schizophrenia (F20), followed by acute polymorphic psychotic disorder without symptoms of schizophrenia (F23) with 13.6%, bipolar disorder (F31) with 10%, and depressive episodes (F32) with 7.3%. When analyzing whether there are differences between the presentation of these diagnoses between the group that presented a first episode vs the group with previous episodes, differences were found regarding the diagnosis of schizophrenia (p< 0.001), occurring mainly in those who had had previous episodes (63.6 %), unlike those who presented a first episode (15.2%). Differences were also found for acute polymorphic psychotic disorder without symptoms of schizophrenia (p < 0.001), presenting mostly in the group with a first episode (42.4%) compared with those who had previous episodes (1.3%, as well as in depressive episodes (p = 0.037), being higher in the group with a first episode (18.2%) vs those who presented previous episodes (3.9%). No statistically significant differences were found in the other diagnoses. The frequencies and percentages of each diagnosis are shown in Table 2.

**Table 2.**
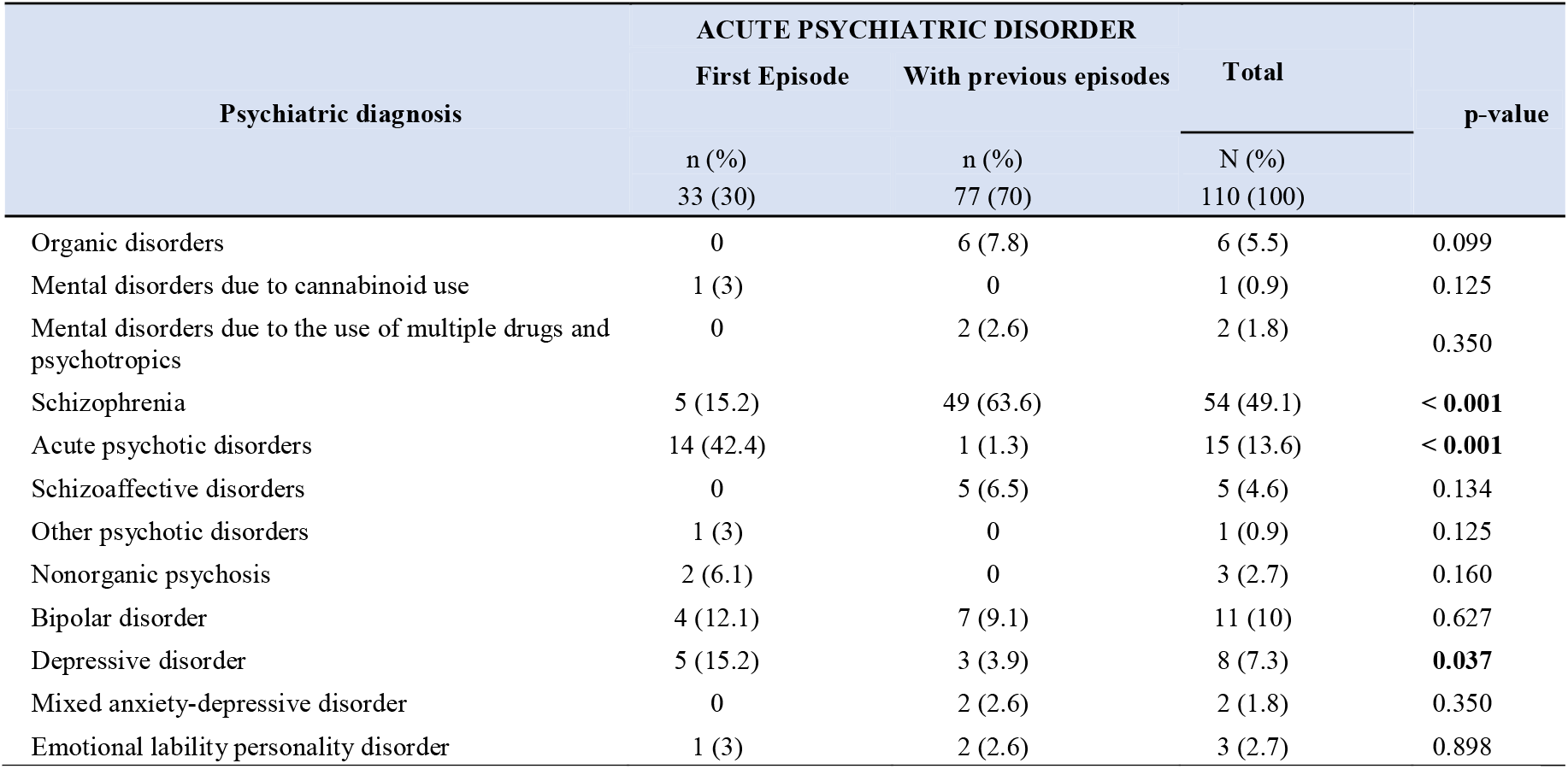
Emergency psychiatric diagnosis according to type of presentation of the acute psychiatric disorder

## DISCUSSION

The presentation of COVID-19 infection is highly heterogeneous and includes neuropsychiatric symptoms ^11^. The psychiatric manifestations associated with COVID-19 may occur due to the direct effect of the virus on the central nervous system, the indirect effect on the immune response, and due to social stressors, which may contribute to the incidence and exacerbation of psychiatric disorders ^18^. An increase in the incidence of the first psychiatric diagnosis has been observed between 14 and 90 days after infection by COVID-19, with a higher risk than for other infections ^11^, this agrees with the results of this study, where - there is a higher frequency of statistically significant cases of acute psychotic disorder as the first episode in patients with COVID-19 infection. In parallel, in a cohort with more than 60,000 cases of COVID-19, during follow-up 3 months after infection, it was observed that 18% of patients were diagnosed with a psychiatric disorder, among which 6% represented a new diagnosis ^19^. In our study, it was observed that 30% of the patients with COVID-19 who attended the emergency department presented a first episode of a psychiatric disorder, a value higher than that reported in the study by *Taquet et al*. ^19^, it is likely that This higher percentage is because the HPHV is a psychiatric reference center, concentrating more cases of patients with psychiatric pathologies.

Approximately 0.9 - 4.0% of people exposed to viral infections develop psychosis, values higher than the incidence in the general population of 15.2 in 100,000 people ^20,21^. In various countries, such as the United States, Spain, Peru, Malaysia, and China, cases of first psychotic episode in patients with COVID-19 have been reported ^3,22–28^. In this study, 42.4% of the patients with the first episode of psychiatric disorders presented an acute psychotic episode. Similarly, an observational study in China reported a 25% increase in the incidence of psychotic disorders, attributed to psychosocial stress associated with the pandemic, although more direct mechanisms are currently being proposed ^29^.The mechanism that produces the first psychotic episode in patients with COVID-19 is unknown, but it is important to consider COVID-19 infection as a cause of new cases of psychotic symptoms ^22^. Patients with psychosis related to COVID-19 may present elevations in C-reactive protein (CRP), ferritin, lactate dehydrogenase (LDH), D-dimer, and increased or decreased levels of leukocytes and platelets ^23,30,31^, which could be considered for future causality studies. Thus far, there is not enough information to clarify the typical presentation of psychosis due to COVID-19, but behavioral disorganization and confusional features are frequently described ^23,25^. compared with patients with psychosis precipitated by the stress of the pandemic, patients with COVID-19 psychosis are less likely to present with COVID-19-related paranoid symptoms or delusions, nor do they have a family history of psychosis ^22^. It is more common for these patients to present an atypical age of onset, subacute presentation, and rapid recovery after treatment with low doses of antipsychotics ^23^, relevant data to be considered in subsequent studies where the characteristics of psychosis could be typified by COVID-19.

Post-COVID-19 follow-up studies have reported that asymptomatic and mild infections may have cognitive impairment, delirium, extreme fatigue, and symptoms associated with mood ^3^. The data from our study suggest a higher frequency of first episode cases of depressive disorders in patients with COVID-19 infection. It has been observed that the symptoms of anxiety and depression are more frequent than anxiety and depression disorders in survivors of COVID-19; however, in a study by *Taquet et al*., a new or recurrent anxiety disorder was reported in 12.8%. and depression in 9% ^3,19^, keeping in relation to what was found in our study. During and after COVID-19 infection, patients are at an increased risk of developing depression and anxiety; even, one month after the infection, between 31% and 38% report depressive symptoms, between 22% and 42% anxiety symptoms, and 20% obsessive-compulsive symptoms ^32–34^. During the hospital stay, 36% of the patients reported anxiety and 29% reported depression; and 2 weeks after infection, the prevalence of anxiety and depression decreased, 9% and 20% respectively; but symptoms of acute stress appeared in 25% ^35^. Symptoms of depression and anxiety after COVID-19 infection are more likely in women, those with infected family members, postinfection physical discomfort, severe infection, elevated inflammatory markers, and previous psychiatric disorders ^33,34,36–38^.

Evidence suggests that psychiatric disorders are a risk factor and, in parallel, a consequence of COVID-19 infection ^3^. In a study by *Czeisler et al*., an increased incidence of psychiatric disorders was described in a population of adults with COVID-19 in the United States; In parallel, he noted that some populations were disproportionately affected, such as young adults, Hispanics, African-Americans, essential workers, unpaid caregivers, and those with preexisting psychiatric disorders ^39^. Within the vulnerable group that are patients with previous psychiatric disorders, according to the results obtained in this study, patients with schizophrenia presented more relapses during the COVID-19 infection compared with those who presented a first episode of schizophrenia. A study carried out in India that included 210 people with severe mental disorder, among which 105 had a diagnosis of schizophrenia, during the quarantine one in 5 patients stopped their medication and 30% presented a worsening of psychiatric symptoms ^40^.

In a study by *Liu et al*. in patients with schizophrenia, patients with suspected COVID-19 infection scored significantly higher in depression, anxiety, perceived stress, and sleep quality. Approximately half of the patients with suspected COVID-19 (52.4%) required an increase in the dose of their medication or the addition of a new psychotropic drug for managing psychiatric symptoms ^41^. It has also been described that patients with schizophrenia have a high probability of mortality from Covid-19, even after adjusting for demographic and medical risk factors ^11^. With all this evidence, it has been suggested that patients with schizophrenia have a higher risk of relapse, considering the emotional distress associated with a vulnerable group during the COVID-19 pandemic, limited access to health care, and the risk of discontinuing medication ^42^. It is hypothesized that some factors could worsen psychosis in patients within the schizophrenia spectrum: fear and stress caused by the pandemic, virus infection, distress and isolation among infected people, and treatment of the disease with corticosteroids and other agents ^43^. In parallel, the social restrictions imposed during the pandemic may have had a negative impact on this population ^44^.

However, it is important to point out that our study has some limitations. A retrospective study was conducted, so it was not possible to evaluate comparative rates between groups with and without COVID-19 infection that would allow us to clarify the differences between said groups. The cases included in the study were those corresponding to COVID-19 patients with mild compromise, since patients with moderate and severe respiratory compromise were mainly cared for in general hospitals due to severity, which could introduce a selection bias. in the study. The sample considered in the group with the first acute episode was smal; therefore, so it was not possible to adjust for possible confounding variables. However, despite these limitations, the study meets the objective set on the analysis of acute episodes of psychiatric disorders in COVID-19 patients during the most critical stage of the pandemic, these data being relevant to our country.

## CONCLUSIONS

The characterization of acute psychiatric disorders in COVID-19 patients showed a predominance of the female sex, mostly young adults, single and without occupation. The use of psychoactive substances was observed, mainly the consumption of alcohol, cocaine, marijuana, and tobacco.

A higher frequency of cases of acute psychotic disorder and depressive disorders was determined as the first episode in patients with COVID-19 infection; however, within the group with previous episodes, greater predominance of patients with acute disorders due to schizophrenia was found.

## Data Availability

All data produced in the present study are available upon reasonable request to the authors

